# Xpert MTB/RIF Ultra is highly sensitive for the diagnosis of tuberculosis lymphadenitis in an HIV-endemic setting

**DOI:** 10.1101/2021.06.11.21258751

**Authors:** Stephanie Minnies, Byron W.P. Reeve, Loren Rockman, Georgina Nyawo, Charissa C. Naidoo, Natasha Kitchin, Cornelia Rautenbach, Colleen A. Wright, Andrew Whitelaw, Pawel Schubert, Robin M. Warren, Grant Theron

## Abstract

**Background:** Tuberculosis lymphadenitis (TBL) is the most common extrapulmonary TB (EPTB) manifestation. Xpert MTB/RIF Ultra (Ultra) is a World Health Organization-endorsed diagnostic test, but performance data for TBL, including on non-invasive specimens, are limited.

**Methods:** Fine needle aspiration biopsies (FNABs) from outpatients (≥18 years) with presumptive TBL (n=135) underwent: 1) routine Xpert (later Ultra once programmatically available), 2) a MGIT960 culture (if Xpert- or Ultra-negative, or rifampicin-resistant), and 3) study Ultra. Concentrated paired urine underwent Ultra. Primary analyses used a microbiological reference standard (MRS).

**Results:** In a head-to-head comparison (n=92) of FNAB study Ultra and Xpert, Ultra had increased sensitivity [91% (95% confidence interval 79, 98) vs. 72% (57, 84); p=0.016] and decreased specificity [76% (61, 87) vs. 93% (82, 99); p=0.020], and detected patients not on treatment. HIV nor alternative reference standards affected sensitivity and specificity. In patients with both routine and study Ultras, the latter detected more cases [+20% (0, 42); p=0.034] and, further indicative of potential laboratory-based room-for-improvement, false-negative study Ultras had more PCR inhibition than true-positives. Study Ultra “false-positives” had less mycobacterial DNA than “true-positives” [trace-positive proportions 59% (13/22) vs. 12% (5/51); p<0.001]. Exclusion or recategorization of “traces” removed potential benefits offered over Xpert. Urine Ultra had low sensitivity [18% (7, 35)].

**Conclusions:** Ultra on FNABs is highly sensitive and detects more TBL than Xpert. Patients with FNAB Ultra-positive “trace” results, most of whom will be culture-negative, may require additional clinical investigation. Urine Ultra could reduce the number of patients needing invasive sampling.

## Background

Tuberculosis (TB) is a leading cause of morbidity and mortality globally. In 2019, extrapulmonary TB (EPTB) represented 16% of new TB cases reported [1] and, in HIV-positive populations, can account up to 50% of all TB [2]. TB lymphadenitis (TBL) accounts for 35% of all EPTB [3, 4]. South Africa, with high TB and HIV burden [1], is particularly affected by EPTB and TBL.

TBL is typically diagnosed by examining fine needle aspiration biopsies (FNABs) from affected lymph nodes. This requires specialised sampling and facilities, and tests have suboptimal sensitivity [5]. One widely-used test is Xpert MTB/RIF (Xpert; Cepheid, USA); a semi-automated real-time PCR that rapidly detects *Mycobacterium tuberculosis* complex (MTBC) DNA and rifampicin resistance [6, 7]. A systematic review and meta-analysis showed heterogeneity in the sensitivity of FNAB Xpert vs. microbiological [83% (95% confidence interval: 71, 91) and composite reference standards [81% (72, 88)] [8]. Specificities were 94% (88, 97) and 99% (95, 100), respectively [8]. Most EPTB diagnostic algorithms recommend culture after a negative Xpert [9], however, this creates delay. Better TBL tests are needed.

One potential test is Xpert MTB/RIF Ultra (Ultra), which offers improved sensitivity over Xpert for pulmonary TB, partly enabled by, in addition to *rpoB*, amplification of multi-copy insertion elements (IS*6110*, IS*1081*) [10]. Data on Ultra for TBL are emerging: one retrospective evaluation tested ten Xpert-negative, culture-positive FNABs and found half to be Ultra-positive [11]; another retrospective evaluation (n=25) reported sensitivity and specificity of 94% (71-77) and 100% (63-72), respectively [12]; and a prospective evaluation (n=73) reported a sensitivity and specificity of 78% (40-97) and 78% (66-87), respectively [13]. No studies included head-to-head Xpert and Ultra data. Additionally, since Ultra’s advent, algorithms for TBL diagnosis remain essentially unchanged from the Xpert era – culture is still recommended in Ultra-negative patients. Whether this is needed or, conversely, if culture is needed to confirm positive Ultra results due to specificity concerns associated with the new trace semi-quantitation category [10, 14], requires investigation.

Lastly, FNABs are rarely collected in primary care; patients are referred to district or tertiary facilities, resulting in care cascade gaps [15]. If an Ultra has high sensitivity and specificity on an easily accessible fluid like urine, the need for invasive sampling could be mitigated; potentially drastically reducing provider and patient economic and time costs. To our knowledge, urine Ultra for TBL is unevaluated.

We evaluated the head-to-head diagnostic accuracy of Xpert and Ultra on FNABs, and Ultra on urine in patients with presumptive TBL in a tertiary hospital setting in an HIV-endemic in South Africa. We hypothesised Ultra would show improved sensitivity compared to Xpert.

## Methods and materials

### Ethics statement

The study was approved by the Stellenbosch University Human Research Ethics Committee and Tygerberg General Hospital (TGH) (both N16/04/050).

### Patient recruitment

135 outpatients (≥18 years) with presumptive TBL (swollen lymph node) undergoing routine referral and investigation at a tertiary referral clinic at TGH in Cape Town, South Africa, were consecutively recruited from 25 January 2017-12 March 2019 and gave FNABs and urine. Patients who received TB treatment ≤60 days prior were excluded.

### Fine needle aspirate collection

FNABs were collected by multiple needle passes using a 23-gauge needle and 10 ml syringe. While the needle was inserted, negative suction with a cutting motion was applied for aspiration. The first two passes were used for routine cytology. From each pass, two slides were prepared: the first airdried for Rapidiff staining and the second spray-fixed for Papanicolaou staining (∼25 μl total volume used per pass) (**Figure 1**). The remaining syringe contents were flushed into 1.5 ml TB transport medium [16]. The third pass (5-50 μl) was collected into 700 μl 5% saline (Ysterplaat Medical Supplies, Cape Town, South Africa).

**Figure 1:**
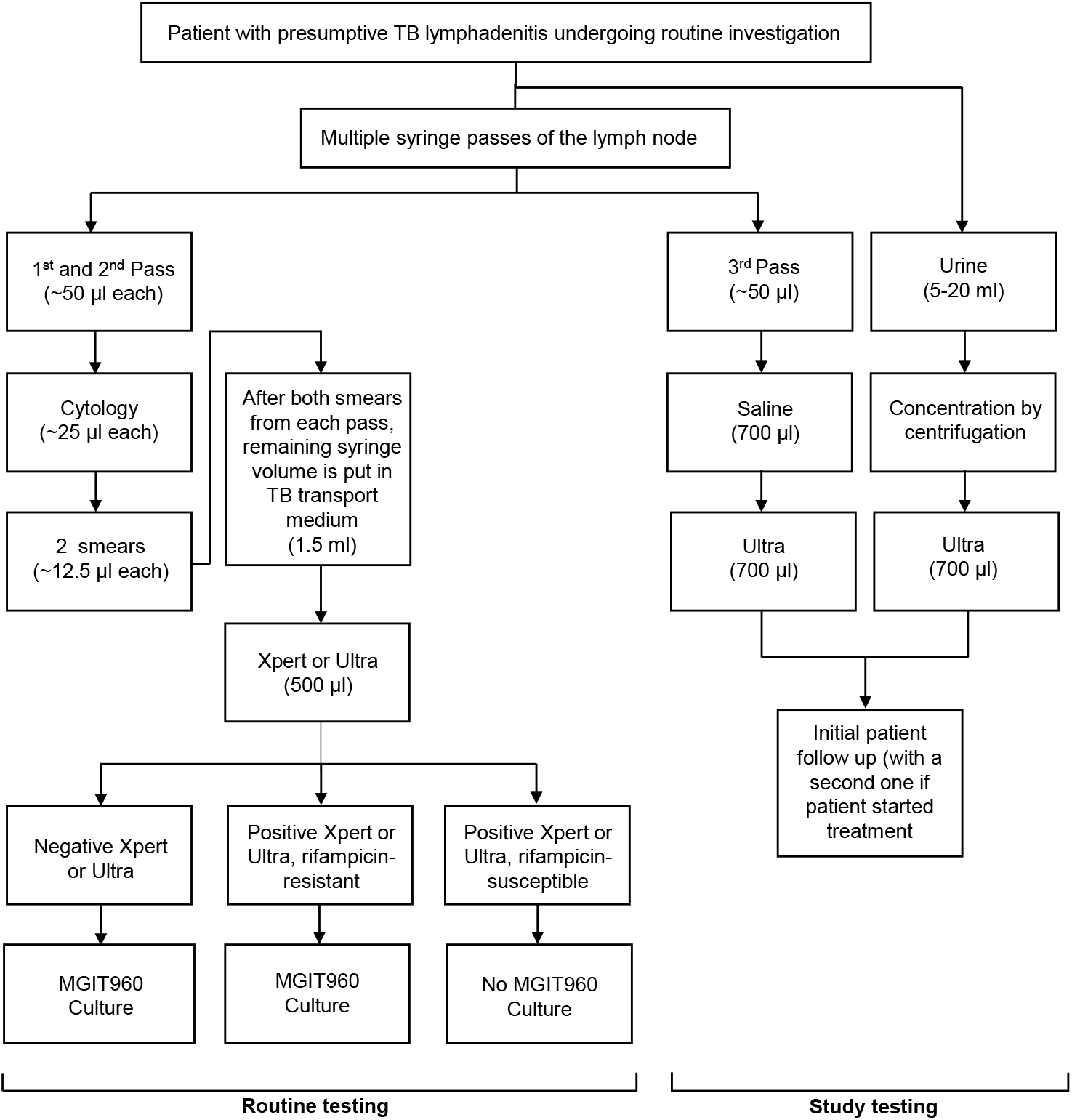
Specimen collection and diagnostic testing in participants with presumptive TB lymphadenitis. Abbreviations: FNAB, fine needle aspirate; TB, tuberculosis; Ultra, Xpert MTB/RIF Ultra; Xpert, Xpert MTB/RIF.

### Xpert, Ultra, and culture

#### Routine testing

Xpert (version 1; Cepheid, USA) was done programmatically from 25 January 2017–9 April 2018 by the government programmatic laboratory [National Health Laboratory Service (NHLS)] who did Ultra (version 1) thereafter [17]. Sample reagent (2 ml; Cepheid, USA) was added to 500 μl of aspirate-containing 1.5 ml TB transport medium (4:1 ratio) and 2 ml of the mixture used for Xpert or Ultra [18, 19]. Per the algorithm, if a specimen was Xpert- or Ultra-positive and rifampicin-susceptible, culture was not done. If Xpert- or Ultra-negative, or Xpert- or Ultra-positive and rifampicin-resistant, 500 μl aspirate-containing TB transport medium was inoculated into a MGIT960 liquid culture without NALC-NaOH decontamination (**Figure 1**). If a non-actionable (not positive or negative) [14] Xpert or Ultra occurred, the remaining 500 μl TB transport medium was used to repeat the test.

#### Study testing

The third pass in 700 μl saline was tested with Ultra (version 3; study Ultra) using a 2:1 sample reagent ratio [19]. Study Ultra was done irrespective of whether routine Xpert or Ultra was done.

#### MTBC typing and drug susceptibility testing

MTBDR*plus* was done on culture-positive isolates for speciation and drug susceptibility testing.

### Urine Ultra

5-20 ml urine stored at −80 °C were centrifuged (1811×*g*, 10 min, room temperature) and the supernatant removed until 700 μl remained, which was tested with Ultra (2:1 sample reagent volume ratio) [19].

### Patient treatment and follow-up

Treatment decisions were programmatic without study involvement (no study results reported for patient management). Attempts were made to telephonically follow-up patients at least 12 weeks after recruitment at which point TB treatment initiation status were recorded and, if treatment started, treatment response was queried. Patients were lost-to-follow-up if at least two calls were unsuccessful, and messages were unreturned for each timepoint.

### Definitions

#### Patient groups

Patients were designated definite, probable, or non-TB using different reference standards. For the microbiological reference standard (MRS), definite TB was culture-positive and/or cytology-positive on FNABs, and non-TBs culture- and cytology-negative on FNABs. Unclassifiable patients had no positive MRS test, culture contaminated or not done, and cytology not done. **Supplementary Table 1** has the extended microbiological standard (eMRS) and composite reference standard (CRS) definitions.

#### Other definitions

Xpert or Ultra actionable results for TB were MTBC-detected and rifampicin-susceptible, rifampicin-resistant or rifampicin-indeterminate, or MTBC not detected [14]. For culture, actionable results were positive or negative for MTBC. For cytology, the presence or absence of granulomatous inflammation was recorded.

### Statistical analysis

We included patients in head-to-head analyses if they had actionable routine index test (Xpert or Ultra), study Ultra, and culture results (or, if culture was non-actionable, a cytology result was available). Proportion tests [20] were done using STATA version 16.0 (StataCorp, College Station Texas, USA) and GraphPad Prism version 8.0.1 (GraphPad Software, San Diego, USA). Venn diagrams were made with InteractiVenn [21]. Differences in diagnostic accuracy metrics were calculated using proportion tests or McNemar’s test as appropriate. STARD guidelines were followed [22].

## Results

### Patient characteristics

Of 135 patients, 44% (59/135) were definite TB and 56% (75/135) non-TBs per the MRS. Characteristics are compared in **Table 1**.

**Table 1:** Demographic and clinical characteristics by microbiological reference standard status. Definite TBs were more likely to be younger, have an involved neck or breast lymph node (vs. another anatomical site) and, if HIV-positive, a lower CD4 count than non-TBs. Data are n (%) or median (IQR).

|  | Overall<br>(n=135) | Definite-TB<br>(n=59) | Non-TB<br>(n=75) |
| --- | --- | --- | --- |
| **Demographics** |  |  |  |
| Age (years) | 36<br>(29-46.5) | 34<br>(27-41) | 39<br>(31.5-47.5)<br>**p=0.019** |
| Female | 72/135<br>(53) | 30/59<br>(51) | 42/75<br>(56)<br>p=0.553 |
| **Clinical characteristics** |  |  |  |
| HIV-positive | 77/133<br>(58) | 35/58<br>(60) | 41/74<br>(55)<br>p=0.569 |
| CD4 count<br>(cells/μl) | 183 (66-304) | 147 (43-281) | 219 (156-358)<br>**p=0.012** |
| Previous TB | 42/135<br>(31) | 19/59<br>(32) | 22/75<br>(29)<br>p=0.720 |
| Pulmonary TB | 38/42<br>(90) | 17/59<br>(29) | 20/75<br>(27)<br>p=0.783 |
| Extrapulmonary<br>TB | 4/42<br>(10) | 2/59<br>(3) | 2/75<br>(3)<br>p=0.807 |
| **Involved site** |  |  |  |
| Neck | 92/134<br>(67) | 53/59<br>(90) | 39/75<br>(52)<br>**p<0.001** |
| Thorax | 16/134<br>(12) | 4/59<br>(7) | 12/75<br>(16)<br>p=0.102 |
| Breast | 9/134<br>(7) | 0/59<br>(0) | 9/75<br>(12)<br>**p=0.006** |
| Other | 17/134<br>(13) | 2/59<br>(3) | 15/75<br>(20)<br>**p=0.004** |

Missing data: HIV, two; CD4, four; lymph node site, one.
One patient was unclassifiable based on case definitions.
“Other” sites included arm (n=3), leg (n=3), groin (n=7), and head (n=4).

### FNAB index test results

76% (103/135) of patients had routine Xpert requested [6/103 (6%) not done] and 24% (32/135) routine Ultra requested [3% (1/32) not done]. Non-actionable results for routine Xpert, routine Ultra and study Ultra were 0% (0/97), 6% (2/31), and 3% (4/135), respectively. 41% (40/97) of routine Xperts were positive (remainder negative). For routine Ultra, 38% (11/29) were positive and, for study Ultra, 74/131 positive (56%; p=0.070 vs. routine Ultra) (**Figure 2**). In a head-to-head comparison of patients with actionable results from each test (study Ultra, routine Xpert, culture, cytology) 37% (22/59), 8% (5/59), 20% (12/59) and 24% (14/59) were positive by each test (**Figure 3A**; study Ultra had the highest yield). 12% (7/59) of these patients with at least one positive result were exclusively detected by study Ultra (cytology exclusively detected two). This proportion detected only by study Ultra (and hence were negative by routine Xpert and/or cytology) increased to 22% (13/59) when culture results, which are not available for rapid clinical decision making, were omitted.

**Figure 2:**
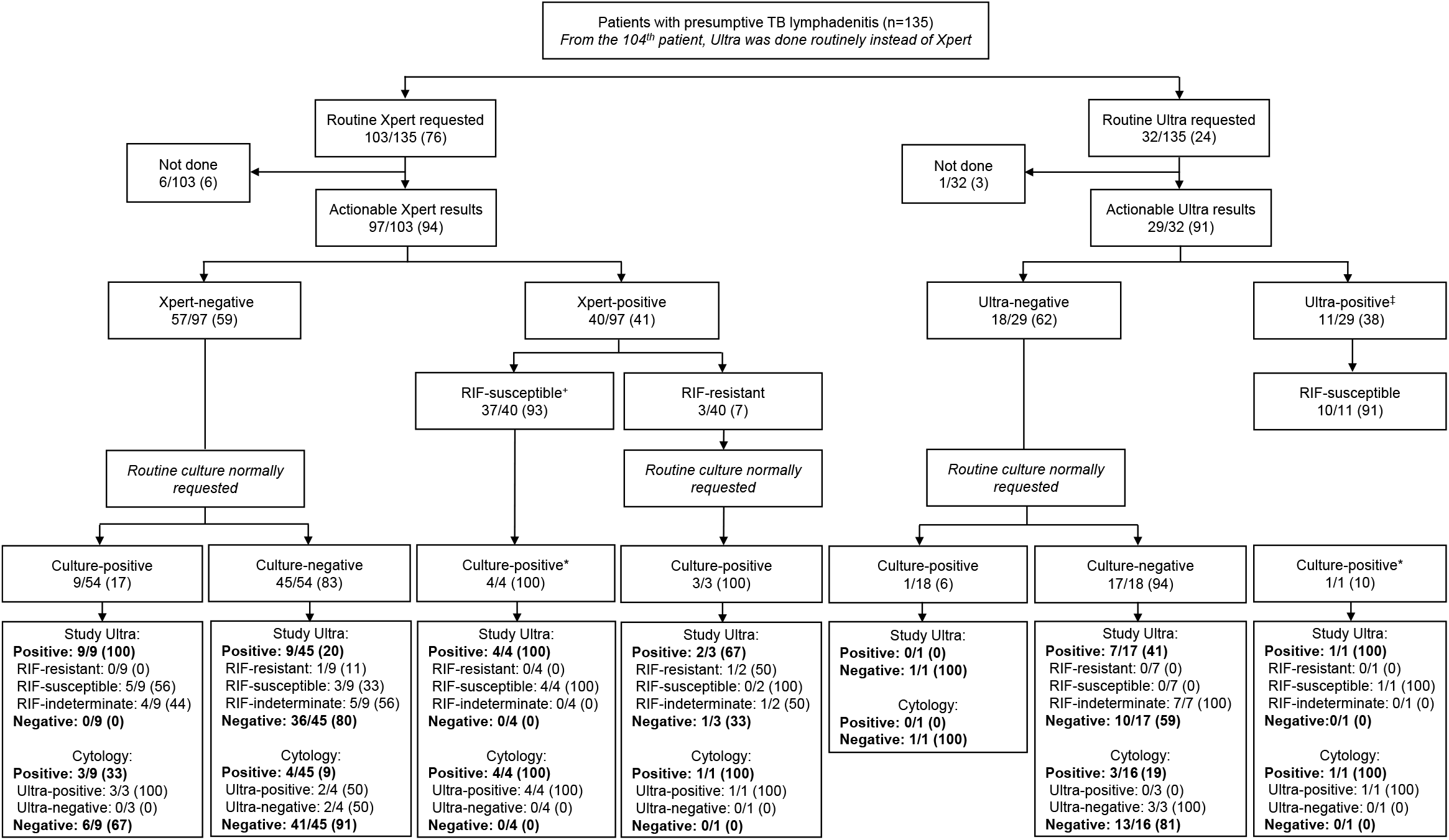
Overview of different FNAB-based test results. Tests done as part of the routine diagnostic algorithm (Xpert later replaced by Ultra, cytology, and culture) and the study (Ultra) are shown. Study Ultra detected TB in most culture-positive FNABs and some culture-negative FNABs. Italicised text indicates programmatic testing (programmatic algorithm adherence imperfect). Data are n/N (%). Abbreviations: RIF, rifampicin; TB, tuberculosis; Ultra, Xpert MTB/RIF Ultra; Xpert, Xpert MTB/RIF. ^+^One routine Xpert-positive, rifampicin (RIF)-susceptible patient had a contaminated culture but was study Ultra-positive, RIF-resistant and 32 routine Xpert-positive, rifampicin (RIF)-susceptible patients had no culture per the **Figure 1** algorithm. ^‡^One routine Ultra was trace-positive RIF-indeterminate. ^*^Culture not normally requested per the routine algorithm. Ultra results under cytology subheadings (in the last row of boxes) are routine not study Ultras. Missing data: In patients with a routine Xpert-negative result, one had a contaminated culture and two were culture not done. Two routine Ultras were non-actionable. Three FNABs did not have cytology done.

**Figure 3:**
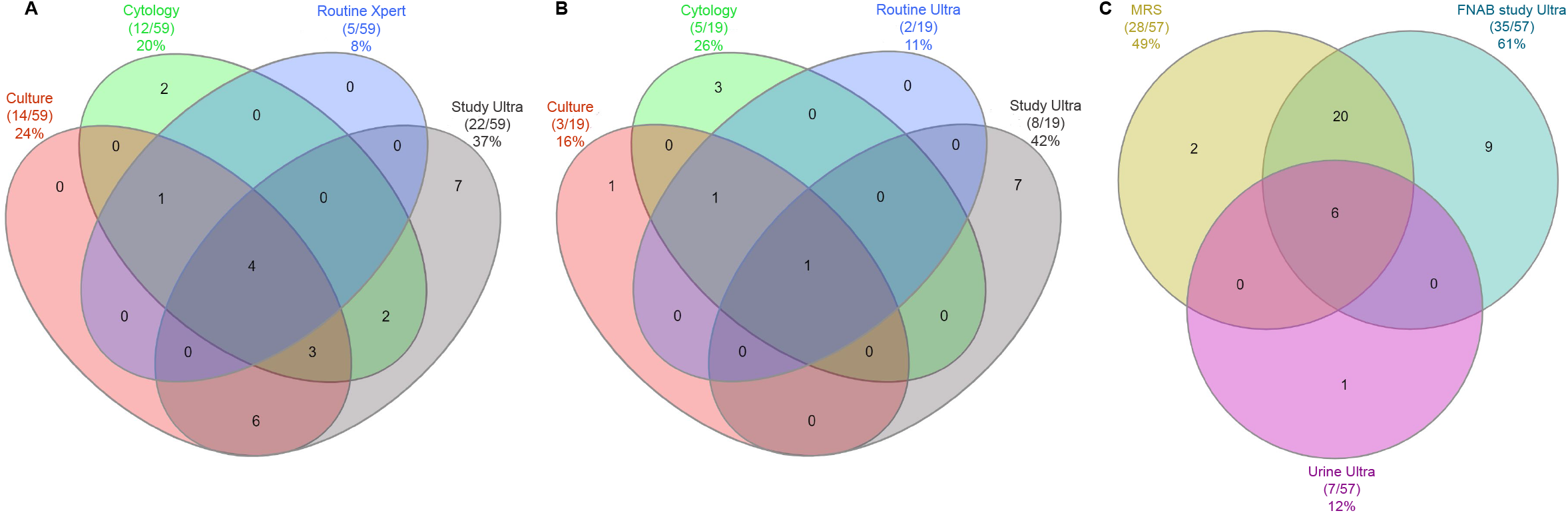
Venn diagrams showing positive results from different FNAB tests (after the 104^th^ participant, Ultra was routinely done instead of Xpert) and urine Ultra. **(A)** Study Ultra, routine Xpert, culture and cytology results in 59 patients. Study Ultra was positive in seven FNABs undetected by routine Xpert. **(B)** Routine Ultra results relative to Study Ultra, routine Ultra, culture, and cytology in 19 patients. Study Ultra was exclusively positive in 36% (7/19) FNABs not detected by routine Ultra, culture and cytology, and had the highest yield. **(C)** Urine Ultra results relative to FNAB study Ultra and the MRS in 57 HIV-positive patients (Urine Ultra negative in all HIV-negatives). Urine Ultra detects less TBL than FNAB study Ultra but could obviate the need for TB diagnostic FNABs in some patients. Data are n/N (%). Abbreviations: FNAB, fine needle aspirate; MRS, microbiological reference standard; TB, tuberculosis; Ultra, Xpert MTB/RIF Ultra; Xpert, Xpert MTB/RIF.

### Diagnostic accuracy and yield of study Ultra and routine Xpert on FNABs

#### Overall

When Ultra was compared head-to-head to Xpert using the MRS (n=92) (**Table 2**), Ultra had improved sensitivity [91% (95% confidence interval: 79, 98) vs. 72% (57, 85); p=0.016] and decreased specificity [76% (61, 87) vs. 93% (82, 99); p=0.020]. Ultra’s positive predictive value (PPV) [79% (66, 89) vs. 92% (78, 98); p=0.114] and negative predictive value (NPV) were like Xpert’s [90% (76, 97) vs. 77% (64, 87); p=0.105]. Conclusions were unchanged for non-head-to-head comparisons, eMRS or CRS (**Table 2, Supplementary results**). Compared to MTBDR*plus* on isolates, no false-negative or false-positive Ultra rifampicin-resistance results occurred, however, numbers were small, precluding precise accuracy estimates (**Supplementary Results**).

**Table 2:** Diagnostic accuracy analyses (non-head-to-head above, head-to-head below) of routine Xpert and study Ultra on FNABs using a MRS (culture and cytology) for *Mycobacterium tuberculosis* complex DNA detection stratified by HIV status. Study Ultra has improved sensitivity compared to routine Xpert but lower specificity. The relative performances of Xpert and Ultra had similar patterns by HIV status and versus the eMRS or CRS (**Supplementary Table 2**). Data are %, 95% CI, and n/N.

|  | Non-head-to-head |  |  |  |  |  |  |  |  |  |  |  |
| --- | --- | --- | --- | --- | --- | --- | --- | --- | --- | --- | --- | --- |
|  | All patients |  |  |  | HIV-negative |  |  |  | HIV-positive |  |  |  |
|  | n=96 |  |  |  | n=36/96 (38) |  |  |  | n=60/96 (62) |  |  |  |
|  | Sensitivity | Specificity | PPV | NPV | Sensitivity | Specificity | PPV | NPV | Sensitivity | Specificity | PPV | NPV |
| Xpert | 73 (58, 85)<br>35/48 | 92 (80, 98)<br>44/48 | 90 (76, 97)<br>35/39 | 77 (64, 87)<br>44/57 | 65 (38, 86)<br>11/17 | 89 (67, 99)<br>17/19 | 85 (55, 98)<br>11/13 | 74 (52, 90)<br>17/23 | 77 (59, 90)<br>24/31<br>p=0.343* | 93 (77, 99)<br>27/29<br>p=0.656* | 92 (75, 99)<br>24/26<br>p=0.455* | 79 (62, 91)<br>27/34<br>p=0.627* |
|  | n=130 |  |  |  | n=55/128 (43) |  |  |  | n=73/128 (47) |  |  |  |
| Ultra | 85 (73, 93)<br>51/60<br>p=0.121‡ | 69 (56, 79)<br>48/70<br>p=0.003‡ | 70 (58, 80)<br>51/73<br>p=0.018‡ | 84 (72, 93)<br>48/57<br>p=0.343‡ | 76 (55, 91)<br>19/25<br>p=0.427‡ | 70 (51, 85)<br>21/30<br>p=0.111‡ | 68 (48, 84)<br>19/28<br>p=0.260‡ | 78 (58, 91)<br>21/27<br>p=0.750‡ | 91 (76, 98)<br>31/34<br>p=0.125‡<br>p=0.109* | 67 (50, 81)<br>26/39<br>p=0.009‡<br>p=0.768* | 70 (55, 83)<br>31/44<br>p=0.031‡<br>p=0.816* | 90 (73, 98)<br>26/29<br>p=0.267‡<br>p=0.227* |
| Δ if traces excluded | -2 (-15, 12)<br>p=0.808§ | +15 (1, 30)<br>p=0.041§ | +13 (-1, 28)<br>p=0.081§ | 0 (-13, 13)<br>p>0.999§ | -3 (-28, 22)<br>p=0.797§ | +21 (1, 41)<br>p=0.076§ | +21 (-2, 44)<br>p=0.103§ | 0 (-22, 22)<br>p>0.999§ | -1 (-15, 13)<br>p=0.905§ | +12 (-8, 32)<br>p=0.253§ | +10 (-9, 28)<br>p=0.332§ | 0 (-16, 16)<br>p>0.999§ |
| Δ if traces reclassified | -10 (-19, -1)<br>p=0.014§ | +18 (8, 29)<br>p<0.001§ | +13 (-1, 28)<br>p=0.081§ | -4 (-17, 9)<br>p=0.558§ | -12 (-29, 5)<br>p=0.083§ | +23 (5, 42)<br>p=0.008§ | +21 (-2, 44)<br>p=0.103§ | -2 (-23, 18)<br>p=0.845§ | -9 (-21, 4)<br>p=0.083§ | +15 (1, 29)<br>p=0.014§ | +10 (-9, 28)<br>p=0.332§ | -6 (-21, 11)<br>p=0.517§ |
|  | Head-to-head |  |  |  |  |  |  |  |  |  |  |  |
|  | n=92 |  |  |  | n=35/92 (38) |  |  |  | n=57/92 (62) |  |  |  |
|  | Sensitivity | Specificity | PPV | NPV | Sensitivity | Specificity | PPV | NPV | Sensitivity | Specificity | PPV | NPV |
| Xpert | 72 (57, 84)<br>33/46 | 93 (82, 99)<br>43/46 | 92 (78, 98)<br>33/36 | 77 (64, 87)<br>43/56 | 65 (38, 86)<br>11/17 | 94 (73, 100)<br>17/18 | 92 (62, 100)<br>11/12 | 74 (52, 90)<br>17/23 | 76 (56, 96)<br>22/29<br>p=0.417* | 93 (76, 99)<br>26/28<br>p=0.832* | 92 (73, 99)<br>22/24<br>p>0.999* | 79 (61, 91)<br>26/33<br>p=0.671* |
| Ultra | 91 (79, 98)<br>42/46<br>p=0.016‡ | 76 (61, 87)<br>35/46<br>p=0.020‡ | 79 (66, 89)<br>42/53<br>p=0.114‡ | 90 (76, 97)<br>35/39<br>p=0.105‡ | 82 (57, 96)<br>14/17<br>p=0.244‡ | 72 (47, 90)<br>13/18<br>p=0.074‡ | 74 (49, 91)<br>14/19<br>p=0.217‡ | 81 (54, 96)<br>13/16<br>p=0.593‡ | 97 (82, 100)<br>28/29<br>p=0.022‡<br>p=0.099* | 79 (59, 92)<br>22/28<br>p=0.127‡<br>p=0.622* | 82 (65, 93)<br>28/34<br>p=0.311‡<br>p=0.455* | 96 (78, 100)<br>22/23<br>p=0.076‡<br>p=0.145* |
| Δ if traces excluded | -1 (-17, 11)<br>p=0.836§ | +7 (-9, 24)<br>p=0.400§ | +5 (-11, 20)<br>p=0.576§ | 0 (-13, 13)<br>p>0.999§ | -3 (-32, 24)<br>p=0.791§ | +14 (-12, 41)<br>p=0.321§ | +11 (-17, 39)<br>p=0.463§ | 0 (-27, 27)<br>p>0.999§ | 1 (-10, 10)<br>p=0.937§ | +3 (-18, 24)<br>p=0.787§ | +1 (-18, 19)<br>p=0.917§ | 0 (-12, 12)<br>p>0.999§ |
| **Δ if traces reclassified** | -13 (-25, 1)<br>**p=0.014<sup>§</sup>** | +9 (-2, 19)<br>**p=0.046<sup>§</sup>** | +5 (-11, 20)<br>p=0.576<sup>§</sup> | -10 (-25, 5)<br>p=0.196<sup>§</sup> | -17 (-42, 6)<br>p=0.083<sup>§</sup> | +17 (-6, 39)<br>p=0.083<sup>§</sup> | +11 (-17, 39)<br>p=0.462<sup>§</sup> | -8 (-35, 18)<br>p=0.542<sup>§</sup> | -11 (-25, 4)<br>p=0.083<sup>§</sup> | +3 (-7, 14)<br>p=0.317<sup>§</sup> | +1 (-18, 19)<br>p=0.917<sup>§</sup> | -11 (-26, 5)<br>p=0.219<sup>§</sup> |

Missing data in the non-head-to-head table: Unclassifiable Ultra, n=1; non-actionable Ultras, n=4; HIV, n=2.
Within column p-values: <sup>‡</sup>Xpert vs. Ultra within an analysis (non-head-to-head, head-to-head) in patients of the same HIV status (overall, negative, positive).
Within row p-values: <sup>\*</sup>HIV-negative vs. HIV-positive within an analysis (non-head-to-head, head-to-head).
Abbreviations: CI, confidence interval; CRS, composite reference standard; eMRS, extended microbiological reference standard; FNABs, fine needle aspirate biopsies; MRS,
microbiological reference standard; NPV, negative predictive value; PPV, positive predictive value; Ultra, Xpert MTB/RIF Ultra; Xpert, Xpert MTB/RIF

#### HIV

Sensitivities and specificities did not differ in HIV-positives vs. -negatives for study Ultra or routine Xpert (**Table 2**). Within HIV-positives, Ultra had improved sensitivity [97% (82, 100) vs. 76% (56, 96); p=0.022] and similar specificity [79% (59, 92) vs. 93% (76, 99); p=0.127] to Xpert.

#### Trace semi-quantitation exclusion or reclassification

When study Ultra traces were excluded, sensitivity [-1% (−17, 11); p=0.836] and specificity [+7% (−9, 24); p=0.400] were unchanged. When trace results were reclassified as negative, sensitivity decreased [-13% (−25, 1), p=0.014] and specificity increased [+9% (−2, 19), p=0.046] (**Table 2**).

#### Ultra PCR inhibition

An analysis of sample processing control (SPC) C_T_ values (**Supplementary Figure 1**; higher values indicate more inhibition) showed more inhibition in study Ultra positives than -negatives [25.80 (IQR: 24.78-27.33) vs. 25.20 (24.55-26.05); p=0.024]. Furthermore, false-negatives were more inhibited than true-positives [26.10 (25.10-28.60) vs. 25.10 (24.00-25.50); p=0.001]; suggesting inhibition contributes to diminished sensitivity.

#### Relationship with bacterial load

Neither Study Ultra nor routine Xpert C_T_ correlated with bacillary load measured using culture time-to-positivity (**Supplementary Figure 2**) in FNABs.

### Comparison of study Ultra true-positive and false-positives

False-positives had less bacterial load than true-positives [IS*6110/*IS*1081* C_T_ 19.00 (IQR: 16.40-21.60) vs. 24.85 (19.88-28.15); p<0.001], a greater proportion were hence “trace” [59% (13/22) vs. 12% (6/51); p<0.001] (**Table 4**). Less inhibition was also observed for the former group [SPC C_T_ 25.05 (24.45-25.95) vs. 26.10 (25.10-28.60); p=0.005]. More study Ultra true-positives were on treatment at follow-up than Ultra false-positives [92% (44/48) vs. 27% (6/22); p<0.001] as more true-positives were positive using a routine test than the false-positives [98% (50/51) vs. 27% (6/22); p<0.001]. The proportions of patients with previous TB in false-vs. true-positives were similar [27% (6/22) vs. 35% (18/51); p=0.503]. The characteristics of true- and false-positives are in **Table 4** and false-positives per patient information in **Supplementary Table 3**.

**Table 3:**
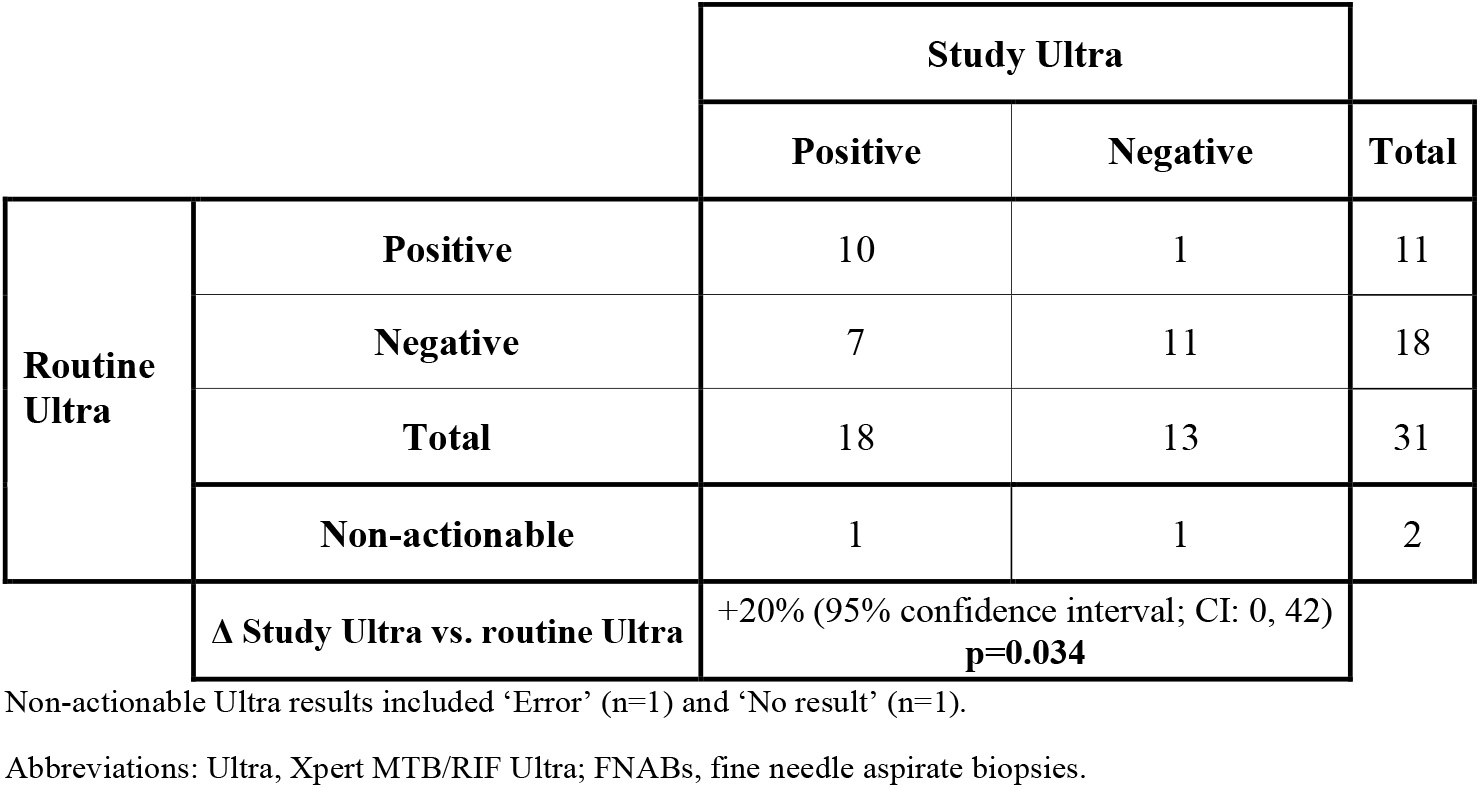
Study and routine Ultra concordance in patients with both tests done on FNABs. More patients were positive by study Ultra (55%) compared to routine Ultra (35%), corresponding to a 20% incremental yield. Study Ultra had no non-actionable results (column not shown).

|  |  | Study Ultra |  |  |
| --- | --- | --- | --- | --- |
|  |  | Positive | Negative | Total |
| Routine Ultra | Positive | 10 | 1 | 11 |
|  | Negative | 7 | 11 | 18 |
|  | Total | 18 | 13 | 31 |
|  | Non-actionable | 1 | 1 | 2 |
| Δ Study Ultra vs. routine Ultra |  | +20% (95% confidence interval; CI: 0, 42)<br>**p=0.034** |  |  |

Non-actionable Ultra results included 'Error' (n=1) and 'No result' (n=1).
Abbreviations: Ultra, Xpert MTB/RIF Ultra; FNABs, fine needle aspirate biopsies.

**Table 4:**
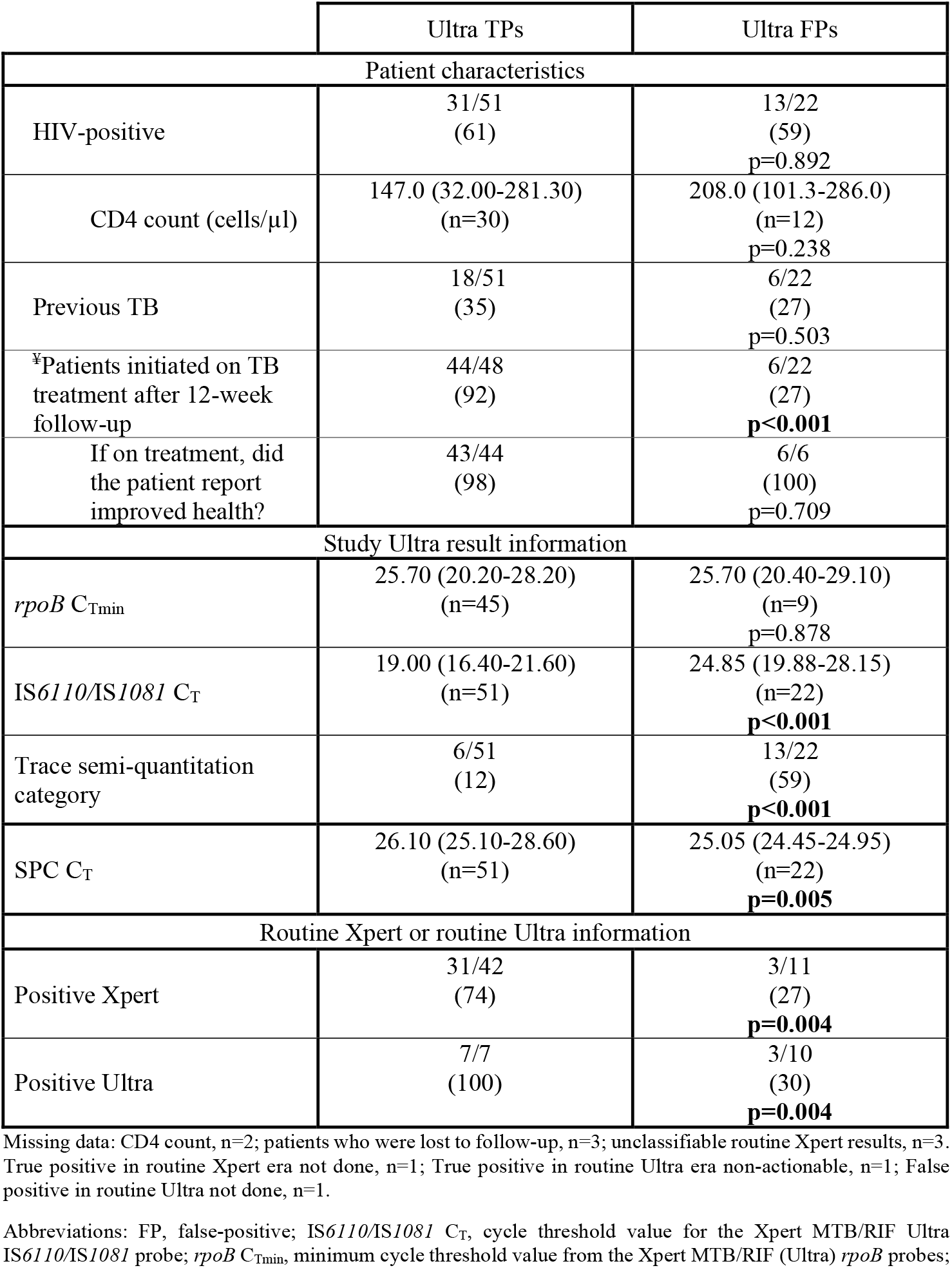
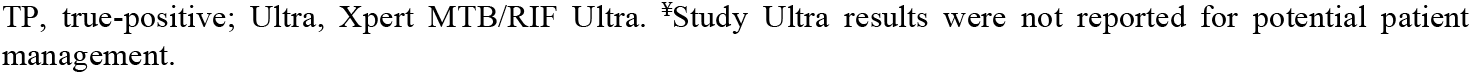
Comparison of patient and microbiology characteristics by whether study Ultra was TP or FP per the MRS. FPs were less likely to have been placed on treatment, had less bacterial load, and were less likely to have been detected by routine Xpert and routine Ultra than TPs. Data are n (%) or median (IQR).

### Study vs. routine Ultra FNAB results

#### Concordance

In patients who received both study and programmatic Ultras, 55% (17/31) were study Ultra-positive and 35% (11/31) routine Ultra-positive. The former detected +20% (95% confidence interval: 0, 42) more TBL (**Table 3**).

#### PCR inhibition

SPC C_T_ analysis showed no difference between study and routine Ultra [25.10 (IQR: 24.35-25.85) vs. 25.50 (24.20-26.50); p=0.081] (**Supplementary Figure 1A**).

### Urine Ultra yield, sensitivity and specificity, and non-actionable results

Urine Ultra had low sensitivity [18% (7, 35)] and high specificity [98% (88, 100)] (head-to-head comparisons with FNAB study Ultra in **Supplementary Table 4**). Of concentrated urines tested with (n=84), 8% (7/84) were non-actionable and 100% (7/7) of these resolved to actionable when unconcentrated urine was tested (one unconcentrated urine was now Ultra-positive). None of the 18 HIV-negative patients had any positive urine Ultra. 12% (7/57) HIV-positives were urine Ultra-positive (six of seven detected by both positive MRS and study Ultra FNAB result; **Figure 3C**). In other words, when urine Ultra was attempted amongst HIV-positives, 11% (7/64, 3 of which were trace) were positive, meaning that universal concentrated urine Ultra testing in HIV-positives with presumptive TBL could reduce the number of FNABs required for TB diagnosis as few are non-actionable.

### Patient treatment status at follow-up

96% (130/135) of patients were followed-up [median (IQR: 37 (16-65) weeks since recruitment] and 52% (68/130) of those had initiated treatment. Of these, 74% (50/68) had been classified as definite TB and 26% (18/68) as non-TB per the MRS. Of the definite TBs, 88% (44/50) were study Ultra-positive whereas, for the non-TBs, 33% (6/18) were study Ultra-positive. Of the remaining study Ultra-positives followed-up, 29% (20/70) were not placed on treatment [in 65% (13/20) of these, study Ultra was the only positive test], indicating potential missed opportunities for treatment initiation. Regarding the clinical status in patients who started treatment, 94% (64/68) reported treatment completion and, of these, 94% (60/64) reported feeling clinically well. 3% (4/130) patients were documented to have died (one of the four had a positive test result that was exclusively study Ultra positive; none of these four were placed treatment).

## Discussion

Our key findings are: 1) study Ultra on FNABs had, compared to Xpert, improved sensitivity and decreased specificity, and outperformed routine Ultra (tests unaffected by HIV and alternative reference standards); 2) approximately 3 in 10 study Ultra-positives had not been placed on treatment, indicating opportunities to improve TBL treatment with Ultra; 3) excluding study Ultra trace results improved specificity (more so than reclassifying to negative) without large sensitivity costs relative to treating Ultra trace results as positive; 4) Urine Ultra had low sensitivity but could reduce the proportion of presumptive TBL patients who require a FNAB in our setting, and 5) Ultra false-negative results are associated with PCR inhibition. These data show high sensitivity of Ultra on FNABs for TBL with the inclusion of trace-positive results (without which sensitivity benefits over Xpert are not seen).

Ultra on FNABs had increased sensitivity than Xpert, suggesting Ultra is rapid initial test for TBL. Ultra did still not detect, however, approximately 1 in 10 TBL cases; indicating a sustained need for more sensitive tests (especially those that use non-invasive specimens) and a continued role for reflex tests for downstream testing of Ultra-negative FNABs. Importantly, like was done previously for Xpert [23], we showed one likely cause of Ultra false-negativity is increased PCR inhibition, suggesting that optimised specimen processing workflows to better remove interfering agents are still needed to boost sensitivity.

Notably, Ultra had suboptimal specificity (two in ten MRS-negative people were study Ultra-positive). One reason may be that culture and cytology have limitations as reference standards for EPTB [8]. Notably, this finding mirrors prior work on TBL that used tissue in addition to fluid biopsies for Ultra, where a specificity of 78% vs. culture was observed [13]. However, when compared to an eMRS including microbiological tests such as FNAB culture as well as culture and Ultra on non-site-of-disease fluids, FNAB Ultra specificity was 100% in that study. In contrast, we applied microbiological tests only to FNABs and did not exhaustively sample anatomical sites [24], which might underestimate specificity.

Ultra false-positivity was more frequent in patients with less mycobacterial DNA and, in contrast to pulmonary TB, FNAB Ultra false-positivity was not associated with prior TB [14]. The true nature of these Ultra “false-positives” in EPTB requires clarification and is an important topic for future research (in our setting, most “false-positive” patients with presumptive pulmonary TB remain well without treatment) [25, 26]. Such “false-positive” results could be caused by *M. tuberculosis* in FNABs that are not culturable using conventional methods like MGIT960. For example, in animal models, *M. tuberculosis* DNA in lymph nodes is detectable during re-activation of TB, despite no pathological evidence of disease and no culturability. *M. tuberculosis* is hypothesised to then disseminate throughout the body from the lymph node [27]. Moreover, we observed no correlation in bacterial load measured using between Ultra and culture, further supporting the presence of *M. tuberculosis* DNA in the absence of culturability.

Critically, if Ultra trace results were excluded or reclassified to elevate specificity, Ultra would lose sensitivity benefits versus Xpert, however, this sensitivity loss was less for the former strategy than the latter; suggesting exclusion is the preferred strategy for handling trace.

When routine and study Ultra concordance were analysed, study Ultras had higher yield. This may be due to specimen processing (e.g., more sample reagent is used for routine Ultras compared to study Ultras) or cartridge version differences but is overall indicative of an area to improve the diagnosis of TBL within the programme.

Few studies examined Ultra on urine [28-30] and none in patients investigated for TBL. Urine Ultra may obviate the need for invasive sampling (and hence referral to a specialised facility, and associated costs and delays). Despite concentration [31], low yield and sensitivity were observed for urine Ultra, suggesting it could marginally reduce FNAB collection (approximately 1/10). Such a strategy is undermined by elevated non-actionable result rates and cost effectiveness, including the number-needed-to-test, would require prospective investigation and modelling, however, we expect the utility of such an approach to be further enhanced with better urine tests [32].

These results have strengths and limitations. Our study was pragmatic and routine culture not always done and, although our MRS included cytology, multiple cultures (including on specimens from other anatomical sites) may improve specificity estimates. Furthermore, multiple FNAB passes were done to obtain adequate volumes that could have introduced sampling variation, however, FNABs were collected using a standardised protocol by a single health worker.

In conclusion, in a routine clinical setting in patients with presumptive TBL, Ultra detects more TBL than Xpert and would result in more people placed on treatment. This is driven by the added benefit of trace results. Furthermore, programmatic Ultra testing can be optimised on the diagnostic laboratory front, as study Ultra had better performance. Urine Ultra could reduce invasive sampling and associate delays but there remains a need for better urine-based tests for TBL. We recommend that a positive FNAB Ultra result be used to initiate treatment, however, patients with a negative Ultra still require confirmatory testing and many patients with a trace-positive result will be culture-negative. Our study supports Ultra’s use for TBL diagnosis.

## Data Availability

Available from the corresponding author.

## Acknowledgments

The authors thank the National Health Laboratory Services, Tygerberg Hospital, Cape Town, South Africa. The authors also thank Ruth Wilson, Lucille Cupido, Roxanne Higgit, Selisha Naidoo and Zaida Palmer. Research reported in this publication was supported by the South African Medical Research Council. The content is the solely the responsibility of the authors and does not necessarily represent the official views of the South African Medical Research Council. The work was funded by the South African Medical Research Council, Stellenbosch University Faculty of Health Sciences, and the National Research Foundation. GT acknowledges funding from the EDCTP2 programme supported by the European Union (grant SF1401, OPTIMAL DIAGNOSIS) and the National Institute of Allergy and Infection Diseases of the National Institutes of Health (U01AI152087). Cepheid donated cartridges but did not have a role in study design or result interpretation.

