## Supplement for "Xpert MTB/RIF Ultra is highly sensitive for the diagnosis of tuberculosis lymphadenitis in an HIV-endemic setting"

**Supplementary Table 5**: Diagnostic accuracy analyses (non-head-to-head top, head-to-head bottom) of routine Xpert and study Ultra (excluding traces) on FNABs using a MRS stratified by HIV status ........ **Error! Bookmark not defined.**

**Supplementary Table 6:** Non-head-to-head and head-to-head diagnostic accuracy analyses of Xpert and Ultra using a microbiological reference standard (MRS), extended microbiological reference standard (eMRS) and composite reference standard (CRS) for the detection of Mycobacterium tuberculosis complex DNA, excluding trace positive Ultra results ........ 12

**Supplementary Table 7**: Diagnostic accuracy analyses (non-head-to-head top, head-to-head bottom) of routine Xpert and study Ultra (reclassifying traces as negative) on FNABs using a MRS stratified by HIV status ........ **Error! Bookmark not defined.**

**Supplementary Table 8:** Non-head-to-head and head-to-head diagnostic accuracy analyses of Xpert and Ultra using a microbiological reference standard (MRS), extended microbiological reference standard (eMRS) and composite reference standard (CRS) for the detection of Mycobacterium tuberculosis complex DNA, reclassifying trace positive Ultra results as negative ........................................................................................................ 14

**Supplementary Table 1:** Reference standard definitions

| | MRS[*] | eMRS[†] | CRS[‡] |
|---|---|---|---|
| | **Site of disease fluid** | | |
| Xpert | ✗ | ✓ | ✓ |
| MGIT960 Culture | ✓ | ✓ | ✓ |
| Cytology | ✓ | ✓ | ✓ |
| | **Non-site-of disease fluid** | | |
| Smear | ✗ | ✓ | ✓ |
| Xpert | ✗ | ✓ | |
| Ultra | ✗ | ✓ | ✓ |
| MGIT960 Culture | ✗ | ✓ | ✓ |
| | **Treatment information** | | |
| TB treatment initiated | ✗ | ✗ | ✓ |
| Response to treatment self-reported by patient | ✗ | ✗ | ✓ |
| | **Case definitions** | | |
| Reference standard **positive (Definite TB cases)** | Any MRS test positive | Any eMRS test positive | Any eMRS test positive/or TB treatment was initiated and response to treatment documented |
| Reference standard **negative (Non-TB patients)** | No MRS test positive | No eMRS test positive | No eMRS test positive and patient not initiated on treatment |
| **Probable TB patients** | N/A | N/A | No eMRS test positive, but treatment initiated |
| **Unclassifiable** | No positive MRS test and site-of-disease fluid culture contaminated or not done | No positive eMRS test and site-of-disease fluid culture contaminated or not done | No positive eMRS test and site-of-disease fluid culture contaminated or not done, or treatment not initiated |

Abbreviations: Composite reference standard, CRS; Extended reference standard, eMRS; MGIT960 culture, Mycobacteria Growth Indicator Tube 960; Microbiological reference standard, MRS; Smear, smear microscopy; Ultra, Xpert MTB/RIF Ultra; Xpert, Xpert MTB/RIF.

### Definitions

#### Microbiological reference standard

For the microbiological reference standard (MRS), a definite TB case was defined as a fine needle aspirate (FNAB) being culture-positive or cytology-positive and a non-TB patient was defined as being FNAB culture and cytology negative. Patients were unclassifiable if they had no positive MRS test and the site-of-disease culture was either contaminated or not done or cytology was not done.

#### Extended microbiological reference standard

For the extended microbiological reference standard (eMRS), a definite TB case was defined as a FNAB or any other body fluid being culture-, smear-, routine Xpert- or Ultra- positive and a non-TB case was defined as FNAB and other body fluids being culture-, smear-, Xpert-and Ultra- negative. Patients were considered unclassifiable if they had no positive eMRS test and the site-of-disease culture was either contaminated or not done.

#### Composite reference standard

For the composite reference standard (CRS), a definite TB case was defined as a FNAB or any other body fluid being culture-, smear-, Xpert- or Ultra- positive or TB treatment was initiated and response to treatment is documented; a probable-TB case was defined as a FNAB or any other body fluid being culture-, smear-, Xpert- or Ultra- positive or the patient being initiated on TB treatment after the 12-week follow up; and a non-TB case was defined as FNAB and other body fluids being culture-, smear-, Xpert-and Ultra- negative, and the patient was not initiated on TB treatment, and the patient was diagnosed with an alternative disease. Patients were unclassifiable if they had no positive eMRS test and the site-of-disease culture was either contaminated or not done, and treatment was not initiated.

### Supplementary Results

Bacterial load in study Ultra and routine Xpert

No correlations were observed between study Ultra quantitation (IS*6110*/IS*1081* $C_T$ and *rpoB* $C_{Tmin}$) and culture time-to-positivity (TTP) and routine Xpert quantitation (*rpoB* $C_{Tmin}$) and culture TTP (**Supplementary Figure 2**).

Drug susceptibility results of study Ultras on FNABs

Of 74 study Ultra-positive patients, 70% (52/74) were rifampicin-susceptible, 4% (3/74) resistant, and 26% (19/74) indeterminate (all trace). In patients who had actionable study Ultra and culture results (n=84), 20% (17/84) had MTBDR*plus* done. Of these, 18% (3/17) were MTBDR*plus* rifampicin-resistant and 82% (14/17) susceptible. 33% (1/3) of these MTBDR*plus* rifampicin-resistant patients were study Ultra rifampicin-resistant (one study Ultra trace-positive, rifampicin indeterminate and the other study Ultra negative), and 57% (8/14) of MTBDR*plus* rifampicin-susceptible patients were study Ultra rifampicin-susceptible (the remaining four were study Ultra trace-positive, rifampicin indeterminate and other remaining two were study Ultra-negative).

**Supplementary Figure 1:** Spaghetti and box and whiskers plots showing FNAB Ultra SPC $C_T$. **(A)** Study SPC $C_T$ vs. routine SPC $C_T$. (**B**) SPC $C_T$ from positive and negative study Ultras. More inhibition was observed in positives. (**C**) SPC $C_T$ in true-positive vs. false-negative study Ultras, showing that greater inhibition is associated with Ultra missing TBL cases. Abbreviations: FNAB, fine needle aspirate biopsy; SPC $C_T$; sample processing control cycle threshold value for the Xpert MTB/RIF Ultra (Ultra) internal positive control which measures PCR inhibition; Ultra, Xpert MTB/RIF Ultra.

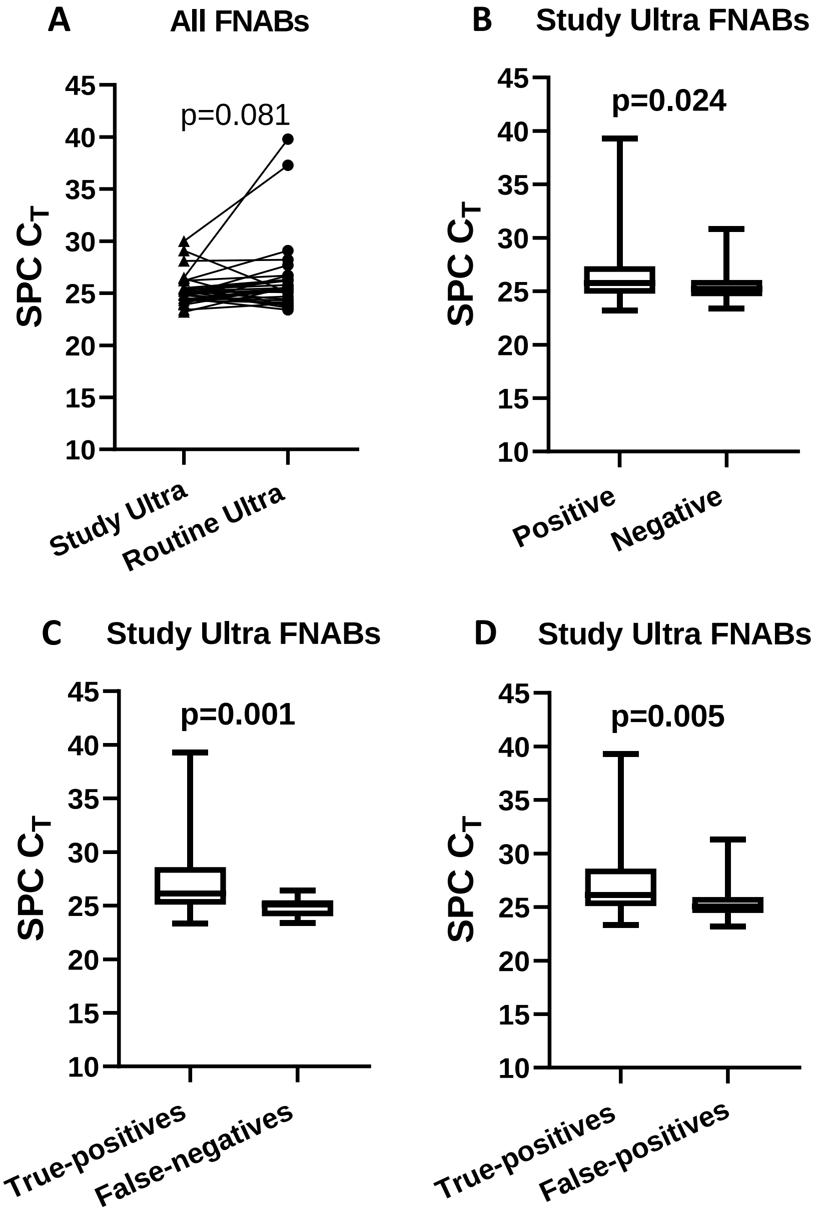

**Supplementary Figure 2:** FNAB Quantitative information of Ultra (IS*6110*/IS*1081*, *rpo*B) and Xpert (*rpo*B) compared with bacillary load (MGIT960 liquid culture TTP)**. (A**) Study Ultra IS*6110*/IS*1081* $C_T$ vs. MGIT960 liquid culture TTP. (B) Study Ultra *rpo*B $C_{Tmin}$ vs. MGIT960 liquid culture TTP**. (C)** Routine Xpert *rpo*B $C_T$min vs. MGIT960 liquid culture TTP. No correlations were observed. Only two culture-positive, routine Ultra-positive FNABs were present and routine Ultra results are hence not graphed. Abbreviations: FNAB, fine needle aspirate biopsy; TTP, culture time-to-positivity; Ultra, Xpert MTB/RIF Ultra; Xpert, Xpert MTB/RIF.

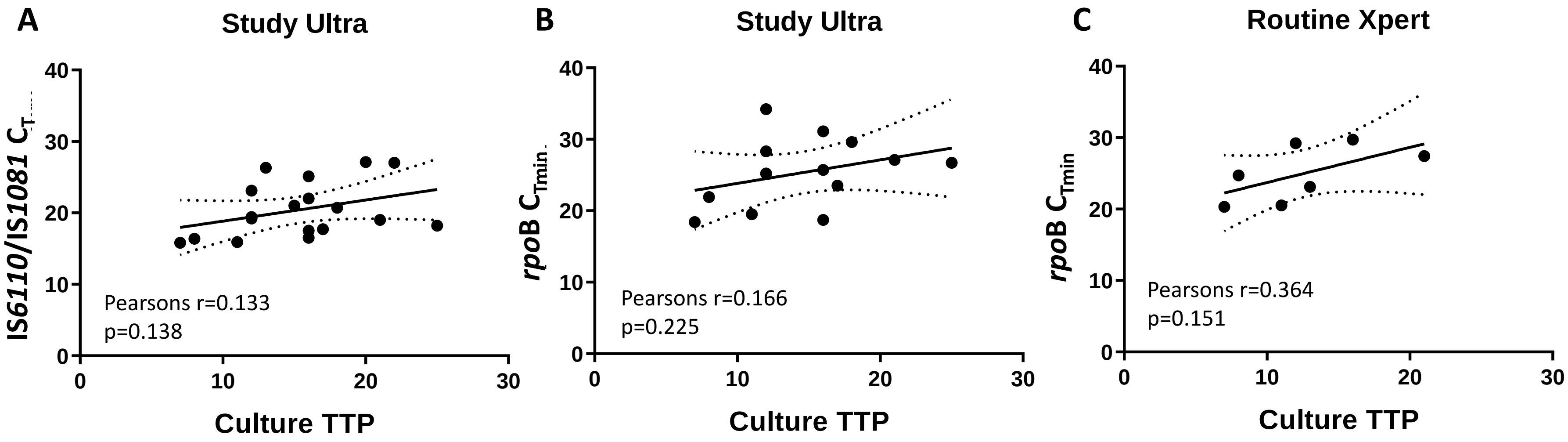

**Supplementary Table 2:** Non-head-to-head and head-to-head diagnostic accuracy analyses of Xpert and Ultra using a MRS, eMRS and CRS for the detection of Mycobacterium tuberculosis complex DNA. Conclusions were like those for the MRS **(Table 2)**. Data are %, 95% CI, and n/N

| | **Non-head-to-head** | | | | | | | | | | | |
|---|---|---|---|---|---|---|---|---|---|---|---|---|
| | **MRS** | | | | **eMRS** | | | | **CRS** | | | |
| | **n=96** | | | | **n=97** | | | | **n=97** | | | |
| | **Sensitivity** | **Specificity** | **PPV** | **NPV** | **Sensitivity** | **Specificity** | **PPV** | **NPV** | **Sensitivity** | **Specificity** | **PPV** | **NPV** |
| **Xpert** | 73 (58, 85)<br>35/48 | 92 (80, 98)<br>44/48 | 90 (76, 97)<br>35/39 | 77 (64, 87)<br>44/57 | 69 (55, 81)<br>36/52<br>p=0.685* | 91 (79, 98)<br>41/45<br>p=0.924* | 90 (76, 97)<br>36/40<br>p=0.970* | 72 (58, 83)<br>41/57<br>p=0.519* | 65 (52, 77)<br>39/60<br>p=0.635±<br>p=0.379¥ | 97 (86, 100)<br>36/37<br>p=0.244±<br>p=0.274¥ | 98 (87, 100)<br>39/40<br>p=0.166±<br>p=0.157¥ | 63 (49, 76)<br>36/57<br>p=0.317±<br>p=0.102¥ |
| | **n=130** | | | | **n=131** | | | | **n=131** | | | |
| **Ultra** | 85 (73, 93)<br>51/60<br>p=0.121‡ | 69 (56, 79)<br>48/70<br>**p=0.003**‡ | 70 (58, 80)<br>51/73<br>**p=0.018**‡ | 84 (76, 97)<br>48/57<br>p=0.343‡ | 83 (71, 91)<br>53/64<br>p=0.085‡<br>p=0.741* | 69 (56, 79)<br>46/67<br>**p=0.005**‡<br>p=0.991* | 72 (60, 81)<br>53/74<br>**p=0.024**‡<br>p=0.815* | 81 (68, 90)<br>46/57<br>p=0.271‡<br>p=0.622* | 76 (65, 85)<br>58/76<br>p=0.147‡<br>p=0.345±<br>p=0.207¥ | 71 (57, 82)<br>39/55<br>**p=0.001**‡<br>p=0.788±<br>p=0.778¥ | 78 (67, 87)<br>58/74<br>**p=0.006**‡<br>p=0.343±<br>p=0.238¥ | 68 (55, 80)<br>39/57<br>p=0.554‡<br>p=0.132±<br>**p=0.047¥** |
| | **Head-to-head** | | | | | | | | | | | |
| | **n=92** | | | | **n=92** | | | | **n=92** | | | |
| **Xpert** | 72 (57, 84)<br>33/46 | 93 (82, 99)<br>43/46 | 92 (78, 98)<br>33/36 | 77 (64, 87)<br>43/56 | 67 (52, 80)<br>33/49<br>p=0.642* | 93 (81, 99)<br>40/43<br>p=0.068* | 92 (78, 98)<br>33/36<br>p>0.999* | 71 (58, 83)<br>40/56<br>p=0.518* | 64 (50, 76)<br>35/55<br>p=0.691±<br>p=0.387¥ | 97 (86, 100)<br>36/37<br>p=0.382±<br>p=0.419¥ | 97 (85, 100)<br>35/36<br>p=0.304±<br>p=0.304¥ | 64 (50, 77)<br>36/56<br>p=0.418±<br>p=0.147¥ |
| **Ultra** | 91 (79, 98)<br>42/46<br>**p=0.016**‡ | 76 (61, 87)<br>35/46<br>**p=0.020**‡ | 79 (66, 89)<br>42/53<br>p=0.114‡ | 90 (76, 97)<br>35/39<br>p=0.105‡ | 88 (75, 95)<br>43/49<br>**p=0.016**‡<br>p=0.573* | 77 (61, 88)<br>33/43<br>**p=0.035**‡<br>p=0.942* | 81 (68, 91)<br>43/53<br>p=0.167‡<br>p=0.808* | 85 (69, 94)<br>33/39<br>p=0.134‡<br>p=0.498* | 84 (71, 92)<br>46/55<br>**p=0.017**‡<br>p=0.551±<br>p=0.252¥ | 81 (65, 92)<br>30/37<br>**p=0.025**‡<br>p=0.636±<br>p=0.583¥ | 87 (75, 95)<br>46/53<br>p=0.091‡<br>p=0.427±<br>p=0.301¥ | 77 (61, 89)<br>30/39<br>p=0.188‡<br>p=0.389±<br>p=0.129¥ |

Within rows: *MRS vs. eMRS, eMRS vs. CRS±, MRS vs. CRS¥; Within columns: Xpert vs. Ultra‡

Abbreviations: CI, confidence interval; CRS, composite reference standard; eMRS, extended microbiological reference standard; MRS, microbiological reference standard; NPV, Negative predictive value; PPV, Positive predictable value; Ultra, Xpert MTB/RIF Ultra; Xpert, Xpert MTB/RIF.

**Supplementary Table 3:** Per patient information for study Ultra-positive patients that were MRS -negative (culture- and cytology-negative) with information on their Ultra semi-quantitation category, previous TB status, TB treatment initiation status and patient's status after at least 12-weeks of follow-up. Data are n/N (%).

| **Patient ID** | **Previous TB** | **Study Ultra semi-quantitation category** | **Routine PCR result** | **Treatment initiated after 12-week follow up** | **Did the patient get better?** (asked telephonically if patient started treatment) |
|---|---|---|---|---|---|
| FNAB038 | No | Trace | Xpert-negative | No | N/A |
| FNAB060 | No | Very Low | Xpert-negative | Yes | Yes |
| FNAB072 | No | Very Low | Xpert-negative | No | N/A |
| FNAB076 | No | Medium | Xpert-positive (Low) | Yes | Yes |
| FNAB110 | No | Trace | Xpert-negative | No | N/A |
| FNAB114 | No | Medium | Xpert-positive (Medium) | No | N/A |
| FNAB128 | No | Trace | Xpert-negative | No | N/A |
| FNAB132 | No | Trace | Xpert-negative | No | N/A |
| FNAB172 | No | Low | Xpert-negative | Yes | Yes |
| FNAB180 | Yes | Low | Xpert-negative | No | N/A |
| FNAB200 | No | Low | Xpert-positive (Low) | Yes | Yes |
| FNAB206 | Yes | Trace | Not done | No | N/A |
| FNAB210 | Yes | Trace | Ultra-negative | No | N/A |
| FNAB214 | No | Trace | Ultra-negative | No | N/A |
| FNAB218 | Yes | Trace | Ultra-negative | No | N/A |
| FNAB220 | No | Trace | Ultra-positive (Very low) | Yes | Yes |
| FNAB230 | No | Trace | Ultra-negative | No | N/A |
| FNAB232 | Yes | Medium | Ultra-positive (Medium) | Yes | Yes |
| FNAB245 | No | Trace | Ultra-negative | No | N/A |
| FNAB251 | Yes | Trace | Ultra-negative | No | N/A |
| FNAB273 | No | Trace | Ultra-negative | No | N/A |
| FNAB403 | No | Medium | Ultra-positive (Medium) | No | N/A |
| **Overall** | 6/22 (27) | Trace: 13/22 (59)<br>Very low: 2/22 (9)<br>Low: 3/22 (14)<br>Medium: 4/22 (18) | Xpert: 11/21 (52)<br>positive, 3/11 (27)<br>negative, 8/11 (73)<br>Ultra: 10/21 (48)<br>positive, 3/10 (30)<br>negative, 7/10 (70) | 6/22 (27) | 6/6 (100) |

Missing data: Routine PCR not done, n=1.

Abbreviations: FNAB, fine needle aspirate biopsy; PCR, polymerase chain reaction; Ultra, Xpert MTB/RIF Ultra.

If the patient was not initiated on TB treatment, N/A was recorded in the last column.

**Supplementary Table 4:** Diagnostic accuracy of Ultra on urine or FNABs measured using the MRS in a head-to-head analysis stratified by HIV status. Urine Ultra had lower sensitivity than FNAB Ultra but increased specificity **(Table 2)**. Data are %, 95% CI, and n/N

| | **All patients** | | | | **HIV-negative** | | | | **HIV-positive** | | | |
|---|---|---|---|---|---|---|---|---|---|---|---|---|
| | **n=76** | | | | **n=18/75 (24)** | | | | **n=57/75 (76)** | | | |
| | **Sensitivity** | **Specificity** | **PPV** | **NPV** | **Sensitivity** | **Specificity** | **PPV** | **NPV** | **Sensitivity** | **Specificity** | **PPV** | **NPV** |
| **Urine-Ultra** | 18 (7, 35)<br>6/33 | 98 (88, 100)<br>42/43 | 86 (42, 100)<br>6/7 | 61 (48, 72)<br>42/69 | 0 (0, 52)<br>0/5 | 100 (75, 100)<br>13/13 | 0/0 | 72 (47, 90)<br>13/18 | 21 (8, 41)<br>6/28<br>p=0.252[*] | 97 (82, 100)<br>28/29<br>p=498[*] | 86 (42, 100)<br>6/7 | 56 (41, 70)<br>28/50<br>p=0.228[*] |
| **Study FNAB-Ultra** | 91 (76, 98)<br>30/33<br>**p<0.001**[‡] | 60 (44, 75)<br>26/43<br>**p<0.001**[‡] | 64 (49, 77)<br>30/47<br>p=0.252[‡] | 90 (73, 98)<br>26/29<br>**p=0.005**[‡] | 80 (28, 99)<br>4/5<br>**p=0.010**[‡] | 38 (14, 68)<br>5/13<br>**p=0.001**[‡] | 33 (10, 65)<br>4/12 | 83 (36, 100)<br>5/6<br>p=0.586[‡] | 93 (76, 99)<br>26/28<br>**p<0.001**[‡]<br>p=0.357[*] | 69 (49, 85)<br>20/29<br>**p=0.005**[‡]<br>p=0.063[*] | 74 (57, 88)<br>26/35<br>p=0.517[‡]<br>**p=0.011**[*] | 91 (71, 99)<br>20/22<br>**p=0.004**[‡]<br>p=0.595[*] |

Missing data: Non-actionable Ultras (n=1), no HIV (n=1) in the head-to-head table.
Within column p-values: [‡] Urine-Ultra vs. FNAB-Ultra
Within row p-values: [*]HIV-negative vs. HIV-positive
Abbreviations: CI, confidence interval; CRS, composite reference standard; eMRS, extended microbiological reference standard; MRS, microbiological reference standard; NPV, negative predictive value; PPV, positive predictive value; Xpert, Xpert MTB/RIF; Ultra, Xpert MTB/RIF Ultra

**Supplementary Table 5:** Non-head-to-head and head-to-head diagnostic accuracy analyses of Xpert and Ultra using a MRS, eMRS and CRS for the detection of *Mycobacterium tuberculosis complex* DNA with and without exclusion of Ultra trace results. Routine Xpert results were compared to study Ultra results. Ultra has similar diagnostic accuracy compared to Xpert after trace positive exclusion. Study Ultra results with trace excluded were like study Ultra results. Similar trends are seen across reference standards. Data are %, 95% CI, and n/N.

| | **Non-head-to-head** | | | | | | | | | | | |
|---|---|---|---|---|---|---|---|---|---|---|---|---|
| | **MRS** | | | | **eMRS** | | | | **CRS** | | | |
| | **n=96** | | | | **n=97** | | | | **n=97** | | | |
| | **Sensitivity** | **Specificity** | **PPV** | **NPV** | **Sensitivity** | **Specificity** | **PPV** | **NPV** | **Sensitivity** | **Specificity** | **PPV** | **NPV** |
| **Xpert[ω]** | 73 (58, 85)<br>35/48 | 92 (80, 98)<br>44/48 | 90 (76, 97)<br>35/39 | 77 (64, 87)<br>44/57 | 69 (55, 81)<br>36/52<br>p=0.685[*] | 91 (79, 98)<br>41/45<br>p=0.924[*] | 90 (76, 97)<br>36/40<br>p=0.510[*] | 72 (58, 83)<br>41/57<br>p=0.519[*] | 65 (52, 77)<br>39/60<br>p=0.635[±]<br>p=0.379[¥] | 97 (86, 100)<br>36/37<br>p=0.244[±]<br>p=0.274[¥] | 98 (87, 100)<br>39/40<br>p=0.166[±]<br>p=0.157[¥] | 63 (49, 76)<br>36/57<br>p=0.317[±]<br>p=0.102[¥] |
| | **n=111** | | | | **n=112** | | | | **n=112** | | | |
| **Ultra excluding trace** | 83 (71, 92)<br>45/54<br>p=0.201[‡] | 84 (72, 93)<br>48/57<br>p=0.248[‡] | 83 (71, 92)<br>45/54<br>p=0.379[‡] | 84 (72, 93)<br>48/57<br>p=0.343[‡] | 81 (69, 90)<br>47/58<br>p=0.151[‡]<br>p=0.751[*] | 85 (73, 93)<br>46/54<br>p=0.368[‡]<br>p=0.887[*] | 85 (73, 94)<br>47/55<br>p=0.510[‡]<br>p=0.760[*] | 81 (68, 90)<br>46/57<br>p=0.271[‡]<br>p=0.622[*] | 74 (62, 84)<br>51/69<br>p=0.272[‡]<br>p=0.341[±]<br>p=0.210[¥] | 91 (78, 97)<br>39/43<br>p=0.224[‡]<br>p=0.413[±]<br>p=0.340[¥] | 93 (82, 98)<br>51/55<br>p=0.304[‡]<br>p=0.221[±]<br>p=0.130[¥] | 68 (55, 80)<br>39/57<br>p=0.554[‡]<br>p=0.132[±]<br>**p=0.047[¥]** |
| **Δ Trace excluded[Φ]** | -2 (-15, 12)<br>p=0.808[§] | +15 (1, 30)<br>**p=0.041[§]** | +13 (-1, 28)<br>p=0.081[§] | 0 (-13, 13)<br>p>0.999[§] | -2 (-15, 12)<br>p=0.799[§] | +16 (2, 31)<br>**p=0.034[§]** | +13 (-0.03, 28)<br>p=0.063[§] | 0 (-14, 14)<br>p>0.999[§] | -2 (-16, 12)<br>p=0.738[§] | +20 (5, 35)<br>**p=0.016[§]** | +15 (3, 26)<br>**p=0.026[§]** | 0 (-17, 17)<br>p>0.999[§] |
| | **Head-to-head** | | | | | | | | | | | |
| | **n=82** | | | | **n=82** | | | | **n=82** | | | |
| **Xpert[ω]** | 80 (64, 91)<br>32/40 | 93 (81, 99)<br>39/42 | 91 (77, 98)<br>32/35 | 83 (69,92)<br>39/47 | 74 (59, 86)<br>32/43<br>p=0.545[*] | 92 (79, 98)<br>36/39<br>p=0.925[*] | 91 (77, 98)<br>32/35<br>p>0.999[*] | 77 (62, 88)<br>36/47<br>p=0.441[*] | 69 (55, 82)<br>34/49<br>p=0.593[±]<br>p=0.255[¥] | 97 (84, 100)<br>32/33<br>p=0.390[±]<br>p=0.431[¥] | 97 (85, 100)<br>34/35<br>p=0.303[±]<br>p=0.303[¥] | 68 (53, 81)<br>32/47<br>p=0.356[±]<br>p=0.093[¥] |
| **Ultra excluding trace** | 90 (76, 97)<br>36/40<br>p=0.210[‡] | 83 (69, 93)<br>35/42<br>p=0.178[‡] | 84 (69, 93)<br>36/43<br>p=0.311[‡] | 90 (76, 97)<br>35/39<br>p=0.367[‡] | 86 (72, 95)<br>37/43<br>p=0.176[‡]<br>p=0.580[*] | 85 (69, 94)<br>33/39<br>p=0.288[‡]<br>p=0.875[*] | 86 (72, 95)<br>37/43<br>p=0.459[‡]<br>p=0.763[*] | 85 (69, 94)<br>33/39<br>p=0.353[‡]<br>p=0.498[*] | 82 (68, 91)<br>40/49<br>p=0.159[‡]<br>p=0.567[±]<br>p=0.266[¥] | 91 (76, 98)<br>30/33<br>p=0.302[‡]<br>p=0.421[±]<br>p=0.338[¥] | 93 (81, 99)<br>40/43<br>p=0.412[‡]<br>p=0.291<br>p=0.178[¥] | 77 (61, 89)<br>30/39<br>p=0.363[‡]<br>p=0.389[±]<br>p=0.129[¥] |
| **Δ Trace excluded[Φ]** | -1 (-14, 11)<br>p=0.836[§] | +7 (-9, 24)<br>p=0.400[§] | +5 (-11, 20)<br>p=0.576[§] | 0 (-13, 13)<br>p>0.999[§] | -2 (-16, 12)<br>p=0.808[§] | +8 (-9, 25)<br>p=0.369[§] | +5 (-10, 20)<br>p=0.521[§] | 0 (-16, 16)<br>p=0.484[§] | -2 (-17, 13)<br>p=0.788[§] | +10 (-6, 26)<br>p=0.241[§] | +6 (-6, 18)<br>p=0.320[§] | 0 (-18, 18)<br>p>0.999[§] |

Within column p-values: [‡]Xpert vs. Ultra within an analysis (non-head-to-head or head-to-head) in patients of the same HIV status (overall, negative, or positive), [§]Study Ultra results (Supplementary Table 2) vs. study Ultra results excluding trace results within an analysis (non-head-to-head or head-to-head) in patients using different reference standards (MRS, eMRS, or CRS).
Within row p-values: [*]MRS vs. eMRS, [±]eMRS vs. CRS, [¥]MRS vs. CRS within an analysis (non-head-to-head or head-to-head).
[ϖ]Although Xpert data are already shown in Supplementary Table 2, small differences in the number of samples included occur in the head-to-head comparison. For the non-head-to-head comparison the Xpert data are identical to that in Supplementary Table 2 but are included here for readability.
[ɸ]This comparison is Ultra with traces excluded vs. Ultra with traces included and considered positive.
Abbreviations: CRS, composite reference standard; eMRS, extended microbiological reference standard; MRS, microbiological reference standard; Ultra, Xpert MTB/RIF Ultra; Xpert, Xpert MTB/RIF.

**Supplementary Table 6:** Non-head-to-head and head-to-head diagnostic accuracy analyses of Xpert and Ultra using a MRS, eMRS and CRS for the detection of Mycobacterium tuberculosis complex DNA, reclassifying trace positive Ultra results as negative. Routine Xpert results were compared to study Ultra results. Ultra has similar diagnostic accuracy compared to Xpert when trace positive results are reclassified as negative. Study Ultra results with trace reclassified had increased sensitivity and decreased specificity compared to normal study Ultra results. Similar trends are seen across reference standards. Data are %, 95% CI, and n/N.

| | **Non-head-to-head** | | | | | | | | | | | |
|---|---|---|---|---|---|---|---|---|---|---|---|---|
| | **MRS** | | | | **eMRS** | | | | **CRS** | | | |
| | **n=96** | | | | **n=97** | | | | **n=97** | | | |
| | **Sensitivity** | **Specificity** | **PPV** | **NPV** | **Sensitivity** | **Specificity** | **PPV** | **NPV** | **Sensitivity** | **Specificity** | **PPV** | **NPV** |
| **Xpert**ω | 73 (58, 85)<br>35/48 | 92 (80, 98)<br>44/48 | 90 (76, 97)<br>35/39 | 77 (64, 87)<br>44/57 | 69 (55, 81)<br>36/52<br>p=0.685* | 91 (79, 98)<br>41/45<br>p=0.924* | 90 (76, 97)<br>36/40<br>p=0.510* | 72 (58, 83)<br>41/57<br>p=0.519* | 65 (52, 77)<br>39/60<br>p=0.635±<br>p=0.379¥ | 97 (86, 100)<br>36/37<br>p=0.244±<br>p=0.274¥ | 98 (87, 100)<br>39/40<br>p=0.166±<br>p=0.157¥ | 63 (49, 76)<br>36/57<br>p=0.317±<br>p=0.102¥ |
| | **n=130** | | | | **n=131** | | | | **n=131** | | | |
| **Ultra with trace reclassified** | 75 (62, 85)<br>45/60<br>p=0.806‡ | 87 (77, 94)<br>61/70<br>p=0.441‡ | 83 (71, 92)<br>45/54<br>p=0.379‡ | 80 (70, 89)<br>61/76<br>p=0.667‡ | 73 (61, 84)<br>47/64<br>p=0.618‡<br>p=0.843* | 88 (78, 95)<br>59/67<br>p=0.609‡<br>p=0.871* | 85 (73, 94)<br>47/55<br>p=0.510‡<br>p=0.760* | 78 (67, 86)<br>59/76<br>p=0.451‡<br>p=0.691* | 67 (55, 77)<br>51/76<br>p=0.797‡<br>p=0.415±<br>p=0.316¥ | 93 (82, 98)<br>51/55<br>p=0.343‡<br>p=0.389±<br>p=0.310¥ | 93 (82, 98)<br>51/55<br>p=0.304‡<br>p=0.221±<br>p=0.130¥ | 67 (55, 77)<br>51/76<br>p=0.636‡<br>p=0.147±<br>p=0.066¥ |
| **Δ Trace reclassified**ϕ | -10 (-19, -1)<br>**p=0.014§** | +18 (8, 29)<br>**p<0.001§** | +13 (-1, 28)<br>p=0.081§ | -4 (-17, 9)<br>p=0.558§ | +10 (-18, 1)<br>**p=0.014§** | +19 (8, 30)<br>**p<0.001§** | +13 (-0.03, 28) p=0.063§ | -3 (-17, 11)<br>p=0.667§ | -9 (-17, 1)<br>**p=0.008§** | +22 (9, 35)<br>**p=0.001§** | +15 (3, 26)<br>**p=0.026§** | +1 (-17, 14)<br>p=0.873§ |
| | **Head-to-head** | | | | | | | | | | | |
| | **n=92** | | | | **n=92** | | | | **n=92** | | | |
| **Xpert**ω | 72 (57, 84)<br>33/46 | 93 (82, 99)<br>43/46 | 92 (78, 98)<br>33/36 | 77 (64, 87)<br>43/56 | 67 (52, 80)<br>33/49<br>p=0.642* | 93 (81, 99)<br>40/43<br>p=0.932* | 92 (78, 98)<br>33/36<br>p>0.999* | 71 (58, 83)<br>40/56<br>p=0.518* | 64 (50, 76)<br>35/55<br>p=0.691±<br>p=0.387¥ | 97 (86, 100)<br>36/37<br>p=0.382±<br>p=0.419¥ | 97 (85, 100)<br>35/36<br>p=0.304±<br>p=0.303¥ | 64 (50, 77)<br>36/56<br>p=0.418±<br>p=0.147¥ |
| **Ultra with trace reclassified** | 78 (64, 89)<br>36/46<br>p=0.470‡ | 85 (71, 94)<br>39/46<br>p=0.180‡ | 84 (69, 93)<br>36/43<br>p=0.290‡ | 80 (66, 90)<br>39/49<br>p=0.729‡ | 76 (61, 87)<br>37/49<br>p=0.371‡<br>p=0.751* | 86 (72, 95)<br>37/43<br>p=0.291‡<br>p=0.763* | 86 (72, 95)<br>37/43<br>p=0.434‡<br>p=0.763* | 76 (61, 87)<br>37/49<br>p=0.637‡<br>p=0.498* | 73 (59, 84)<br>40/55<br>p=0.306‡<br>p=0.747±<br>p=0.521¥ | 92 (78, 98)<br>34/37<br>p=0.304‡<br>p=0.409±<br>p=0.323¥ | 93 (81, 99)<br>40/43<br>p=0.397‡<br>p=0.291±<br>p=0.178¥ | 69 (55, 82)<br>34/49<br>p=0.580‡<br>p=0.489±<br>p=0.247¥ |
| **Δ Trace reclassified**ϕ | -13 (-25, 1)<br>**p=0.014§** | +9 (-2, 19)<br>**p=0.046§** | +5 (-11, 20)<br>p=0.576§ | -10 (-25, 5)<br>p=0.196§ | -12 (-23, -1)<br>**p=0.014§** | +9 (-2, 20)<br>**p=0.046§** | +5 (-10, 20)<br>p=0.521§ | -9 (-26, 7)<br>p=0.293§ | -11 (-21, -1)<br>**p=0.014§** | +11 (-2, 24)<br>**p=0.046§** | +6 (-6, 18)<br>p=0.320§ | -8 (-26, 11)<br>p=0.430§ |

Within column p-values: [‡]Xpert vs. Ultra within an analysis (non-head-to-head or head-to-head) in patients of the same HIV status (overall, negative, or positive), [§]Study Ultra results (Supplementary Table 2) vs. study Ultra results excluding trace results within an analysis (non-head-to-head or head-to-head) in patients using different reference standards (MRS, eMRS, or CRS).

Within row p-values: [*]MRS vs. eMRS, [±]eMRS vs. CRS, [¥]MRS vs. CRS within an analysis (non-head-to-head or head-to-head).

[ϖ]Although Xpert data are already shown in Supplementary Table 2, small differences in the number of samples included occur in the head-to-head comparison. For the non-head-to-head comparison the Xpert data are identical to that in Supplementary Table 2 but are included here for readability.

[ϕ]This comparison is Ultra with traces reclassified as negative vs. Ultra with traces considered positive.

Abbreviations: CRS, composite reference standard; eMRS, extended microbiological reference standard; MRS, microbiological reference standard; Ultra, Xpert MTB/RIF Ultra; Xpert, Xpert MTB/RIF.
